# A roadmap for implementation of kV-CBCT online adaptive radiation therapy and initial first year experiences

**DOI:** 10.1101/2022.10.03.22280665

**Authors:** Dennis N. Stanley, Joseph Harms, Joel A. Pogue, Jean-Guy Belliveau, Samuel R. Marcrom, Andrew M. McDonald, Michael C. Dobelbower, Drexell H. Boggs, Michael H. Soike, John A. Fiveash, Richard A. Popple, Carlos E. Cardenas

## Abstract

**Purpose:** Online Adaptive Radiation Therapy(oART) follows a different treatment paradigm than conventional radiotherapy and, because of this, the resources, implementation, and workflows needed are unique. The purpose of this report is to outline our institution’s experience establishing, organizing, and implementing an oART program using the Ethos therapy system.

**Methods:** We include resources used; operational models utilized, program creation timelines, and our institutional experiences with implementation and operation of an oART program. Additionally, we provide a detailed summary of our first year’s clinical experience where we delivered over 1000 daily adaptive fractions. For all treatments, the different stages of online adaption, primary patient set-up, initial kV-CBCT acquisition, contouring review and edit of influencer structures, target review and edits, plan evaluation and selection, Mobius3D 2^nd^ check and adaptive QA, 2^nd^ kV-CBCT for positional verification, treatment delivery, and patient leaving the room, were analyzed.

**Results:** We retrospectively analyzed data from ninety-seven patients treated from August 2021-August 2022. 1677 individual fractions were treated and analyzed, 632(38%) were non-adaptive and 1045(62%) were adaptive. 74 of the 97 patients (76%) were treated with standard fractionation and 23 (24%) received stereotactic treatments. For the adaptive treatments, the generated adaptive plan was selected in 92% of treatments. On average(±std), adaptive sessions took 34.52±11.42 minutes from start to finish. The entire adaptive process (from start of contour generation to verification CBCT), performed by the physicist (and physician on select days), was 19.84±8.21 minutes.

**Conclusion:** We present our institution’s experience commissioning an oART program using the Ethos therapy system. It took us 12 months from project inception to treatment of our first patient and 12 months to treat 1000 adaptive fractions. Retrospective analysis of delivered fractions showed that average overall treatment time was approximately 35 minutes and average time for the adaptive component of treatment was approximately 20 minutes.

## 1. Introduction

Since first proposed as a concept in the 1990s[1], adaptive radiation therapy has seen tremendous advances through research and clinical implementation. Several research studies have highlighted the potential benefits of adapting radiation therapy for both accounting for tumor changes throughout the course of treatment and its ability to account for inter-fraction motion [1-6]. Technological advances including, increased image quality, faster cone beam CT (CBCT) acquisition times, iterative reconstruction techniques, GPU based optimization and automatic contouring algorithms have brought about the clinical realization of adaptive radiotherapy, significantly reducing the time required to create new radiotherapy treatment plans that meet standard clinical quality standards.

While few online adaptive radiation therapy (oART) treatment delivery systems have been released for clinical use over the past decade, there has been tremendous increase in the clinical applicability of these systems [5, 7-14]. With oART, clinical teams now have the ability to create custom radiation therapy plans based on the patient’s daily anatomy, rather than the patient’s anatomy from treatment simulation images, which may have been acquired days or weeks before treatment commencement [3]. While this new treatment delivery approach offers the potential to reduce the dose to healthy tissues, improve target localization, and increase tumor control, the technical, administrative, and implementation challenges associated with oART are significant [3, 15-17].

As oART programs are being widely adopted in clinics, it is imperative to anticipate the time and resources that are required with this treatment prior to starting therapy. There is little published data on how much treatment time is needed for delivery of online-adaptive treatments. The need for efficient and streamlined workflow is imperative because increased table times may result in changes to internal anatomy, which can negate the benefits of an adaptive treatment session and require the process to be restarted.

oART follows a different treatment paradigm than conventional radiotherapy and, because of this, the resources, implementation, and workflows needed are unique. The primary purpose of this report is to outline our institution’s experience commissioning an oART program using the Ethos therapy system (Varian Medical Systems, Palo Alto, CA). This report will highlight the resources used, operational models utilized, and program creation timelines. As a secondary endpoint, we include a detailed summary of our clinical experience over one year after releasing the Ethos system for clinical use.

## 2. Methods and Materials

### 2.1. Treatment Machine Description

The Ethos is a 6MV Flattening Filter Free (6X-FFF) ring-based linear accelerator capable of performing intensity-modulated radiation therapy (IMRT) and volumetric modulated arc therapy (VMAT) with an output rate of up to 800 cGy/min at Dmax. It has a treatment gantry speed of up to four rotations per minute (4 RPM) and a dual stacked, 28 cm^2^ × 28 cm^2^ at maximum, multileaf collimator with maximum leaf speeds of 5.0 cm/sec. The Ethos is equipped with MV and kV imagers inside of an enclosed 100 cm wide bore. For the Ethos, only the kV-CBCT can be used during clinical operation while other imaging modalities can be used in service and QA modes. The Ethos has a magnetically driven couch with a 226 kg weight limit and 41.6 cm, 47.5 cm and 165.5 cm of travel in the lateral, vertical and longitudinal directions respectively. The treatment planning system (TPS) for Ethos utilizes vendor-provided golden beam data.

### 2.2. Ethos OART Workflow

The Ethos OART workflow has been elaborated on in detail in a previous publication (cite ref 31). Here we provide a simple overview of the steps involved in an adaptive session. More details are provided in the results section. The adaptive portion of the treatment delivery process consists of three modules: Influencer review, target review, and plan selection. Ethos relies on disease-specific automatic segmentation models as the basis for adaptive treatment planning. Thus, in the treatment planning system, different disease sites are organized into modules, which are called “Intents”. Each Intent is associated with selected organs that are automatically contoured (either using AI-based contouring or deformable image registration-based contouring); these are referred to as the “Influencer Structures” for the intent. Based on these influencers, a structure-guided deformable registration is applied from the planning CT to the CBCT. The deformed planning CT provides Hounsfield units from the simulation CT that deformably reflect the internal anatomy of the day as shown on the planning CBCT. This process results in the generation of a synthetic CT dataset, which is later used, for dose calculation. After influencer review is completed, target volumes are propagated onto the Planning CBCT for review. In the last step, Ethos generates two treatment plans: scheduled and adaptive. The scheduled plan provided is the initially-approved plan that is re-calculated on the synthetic CT. The adaptive plan is a newly optimized and generated plan based on the clinical priorities specified during planning. In this review workspace, a score card is created and the adaptor determines which plan is used for treatment that day.

### 2.3. Retrospective review of first twelve months of Ethos adapted treatment deliveries

To provide more detail on each step in the OART treatment process, we conducted a single institution comprehensive time analysis of the over 1600 fractions delivered over a 12-month period. For all treatments, a machine data log is updated with each intervention or logic decision as entered on the treatment console from the time the patient enters to the room until the session is completed and closed. These log files include timestamps, treatment decisions, plan selections and plan type for each step in the oART treatment delivery workflow. Log files were analyzed for all individual patient’s treatment sessions utilizing custom scripts. Any pre-treatment preparation (e.g. bladder filling or consultation) were not included in the evaluated treatment time. For adaptive patients, the different stages of online adaption were recorded from the patient logs and are as follows: primary patient set-up, initial kV-CBCT acquisition, contouring review and edit of influencer structures, target review and edits, plan evaluation and selection, Mobius3D 2^nd^ check and adaptive QA, 2^nd^ kV-CBCT for positional verification, treatment delivery, and patient leaving the room. Prior to delivery monitor units per beam, total monitor units, target coverage, ROI statistics and 3D gamma were analyzed in Mobius3D adapt, evaluated, and verified by the adaptive physicist.

## 3. Results

### 3.1. Initial implementation

#### 3.1.1. Implementation timeline

Figure 1 shows an overview of our institutional timeline for the implementation of an oART program with the significant milestones denoted. Not all aspects of the project are shown in Figure 1 but the significant preparation and treatment milestones are detailed. Overall, pre-installation planning and training was completed in approximately 6 months. It took approximately 9 months from clinical release to administer 1000 treatments, and approximately one year to deliver 1000 online-adaptive treatments.

**Figure 1:**
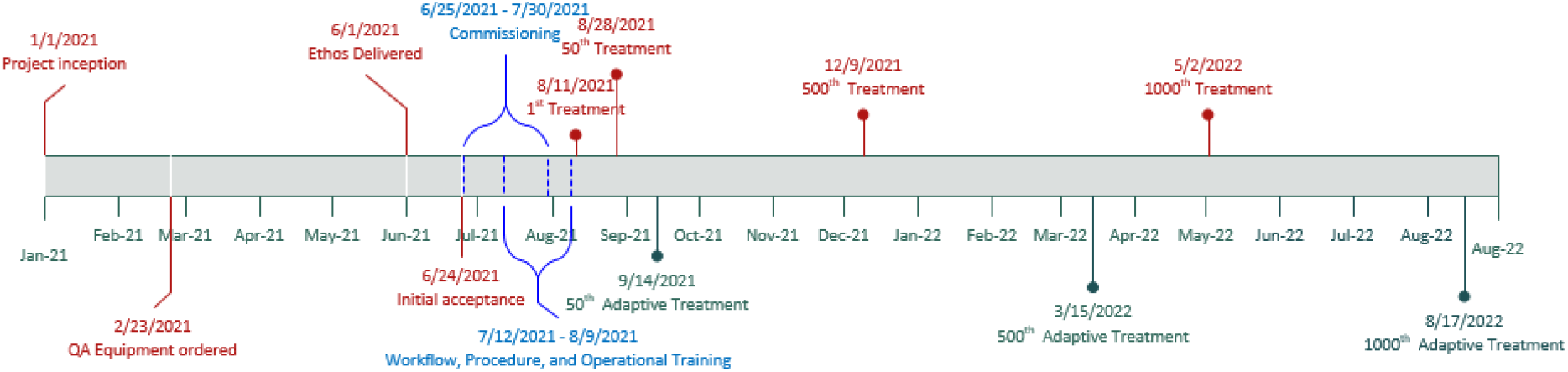
Institutional timeline for the implementation of an oART program and significant clinical milestones after clinical release.

#### 3.1.2. Equipment

Table 1 provides a list of equipment and the primary role of that equipment used in the implementation, commissioning and ongoing QA of a kV-CBCT guided oART program. This list is not a recommendation for specific products or an exhaustive recommendation, rather it was conceived by our clinical team in order to be compliant with standard quality assurance procedures recommended by the American Association of Physicists in Medicine (AAPM) Task Groups 142[18] and 51[19]. All new equipment was ordered approximately 4 months prior to the estimated machine installation to allow for commissioning of the QA devices and cross-calibration with existing equipment. There is some overlap between the devices needed for quality assurance of the oART machine and a conventional C-arm linear accelerator. However, due to the physical limitations of ring gantry accelerators and software limitations of the Ethos treatment planning system, there is increased need to verify device compatibility. For example, only select 3D water tanks will physically fit into the 100 cm bore due to the physical dimensions of the device and inability to remove the couch from inside the bore. Additionally, special considerations for the resources required for training, dosimetry validation, and ongoing quality control should be evaluated. Phantoms were chosen deliberately for the ring gantry environment with special considerations made for ongoing QA workflows and ability to fit inside the specific geometry. A 1D water tank was used for absolute calibration and measurement of depth dose curves and a 2D array was used for verification of beam profiles and off-axis factors. The behavior of the 2D array for profile measurement was previously benchmarked against measurements taken in water on a separate C-arm linear accelerator within our department. Additionally, the Ethos system can only be used with precalibrated “golden beam” data, making verification of the preset profiles with only a 2D array feasible in accordance with the requirements of AAPM’s Medical Physics Practice Guideline 5a.[20].

**Table 1:**
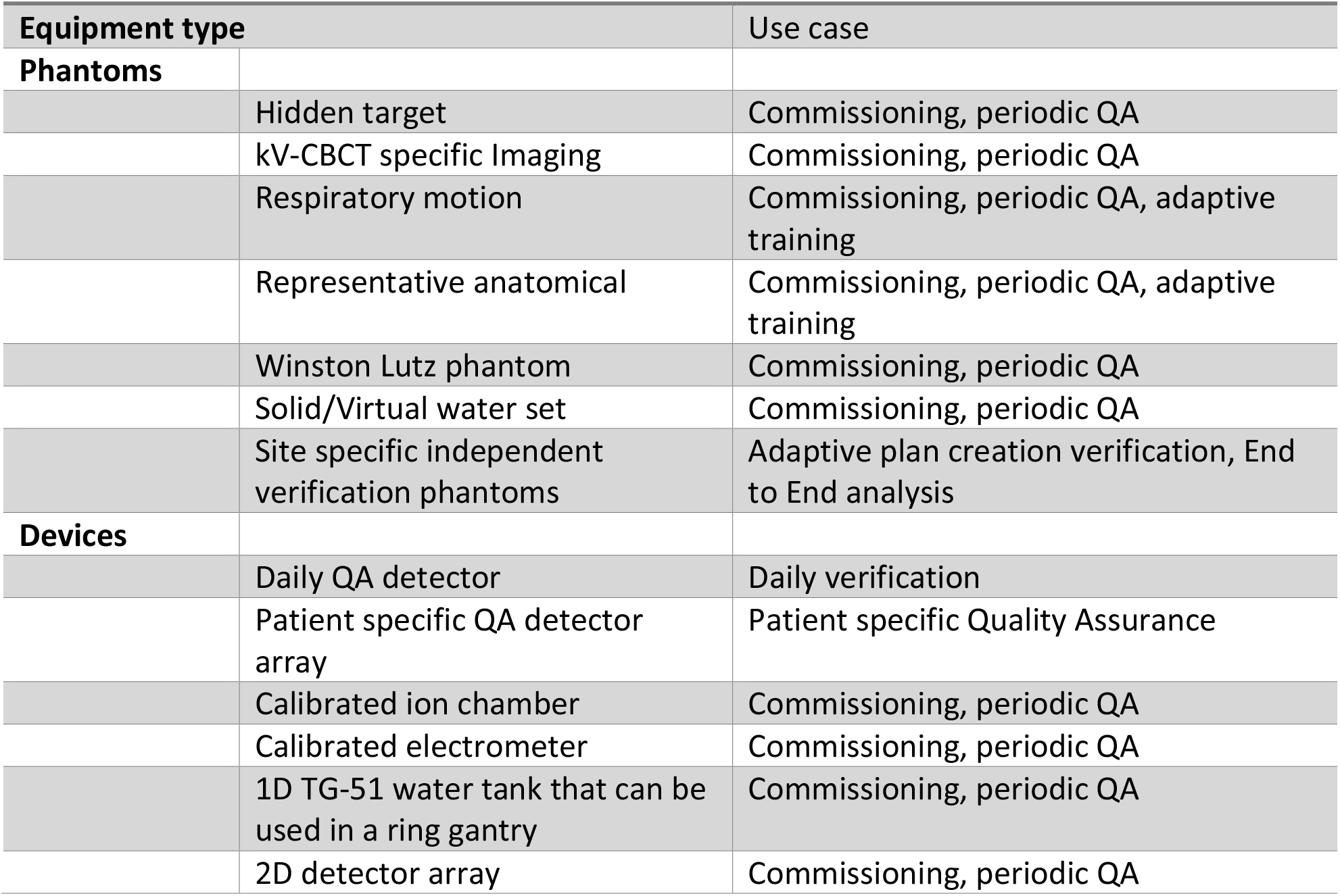
Equipment list used for initial implementation and ongoing QA of an oART program

| Equipment type |  | Use case |
| --- | --- | --- |
| **Phantoms** |  |  |
|  | Hidden target | Commissioning, periodic QA |
|  | kV-CBCT specific Imaging | Commissioning, periodic QA |
|  | Respiratory motion | Commissioning, periodic QA, adaptive training |
|  | Representative anatomical | Commissioning, periodic QA, adaptive training |
|  | Winston Lutz phantom | Commissioning, periodic QA |
|  | Solid/Virtual water set | Commissioning, periodic QA |
|  | Site specific independent verification phantoms | Adaptive plan creation verification, End to End analysis |
| **Devices** |  |  |
|  | Daily QA detector | Daily verification |
|  | Patient specific QA detector array | Patient specific Quality Assurance |
|  | Calibrated ion chamber | Commissioning, periodic QA |
|  | Calibrated electrometer | Commissioning, periodic QA |
|  | 1D TG-51 water tank that can be used in a ring gantry | Commissioning, periodic QA |
|  | 2D detector array | Commissioning, periodic QA |

#### 3.1.3. Commissioning

Commissioning an oART program has three main components: machine, software, and adaptive validation. Netherton et al. [21, 22] provide guidance on a multi-institutional experience of commissioning the Varian Halcyon. Mechanically, the Varian Ethos is very similar to a Varian Halcyon with differences regarding available software and specific machine features. Validation and commissioning of the Ethos treatment planning system was done following previously published guidance [18-20, 22-24] and in accordance with standard practice. Due to the novel nature of online adaptive planning with real time plan creation, we placed an increased emphasis on disease site-specific, independent end-to-end verification in an adaptive environment. Additionally, we modified some in-house phantoms and software to address specific aspects of the adaptive process that we felt were not adequately addressed by conventional phantoms. For example, to simulate anatomical changes between fractions, conventional phantoms were scanned with layers of bolus ranging from 0 cm to 5 cm. Using these scans, simulations of adaptive treatments were performed in an Ethos treatment emulator. The Ethos treatment emulator is a virtual sandbox system provided by Varian for initial testing and configuration. It should be noted that this is not an exhaustive list of every test performed during commissioning and acceptance and individuals should refer to the vendor specific and task group recommended tests for guidance [18-25]. Since the Varian Ethos comes with an un-editable, preinstalled beam model, traditional commissioning becomes more of a verification task and fewer measurements and adjustments may be required than traditional linear accelerators. Table 2 shows a summary of selected results from commissioning with an estimated time for completion.

**Table 2:**
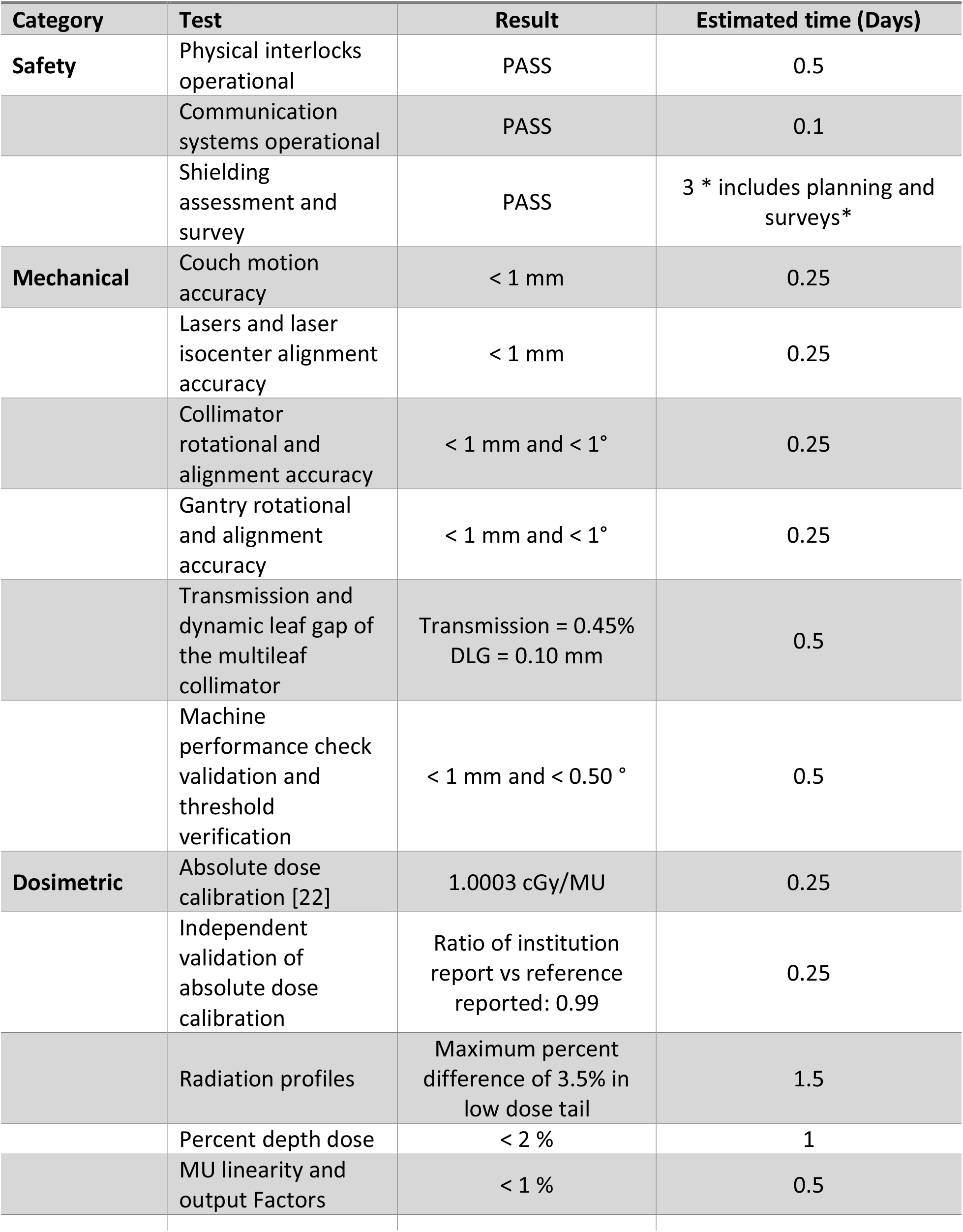
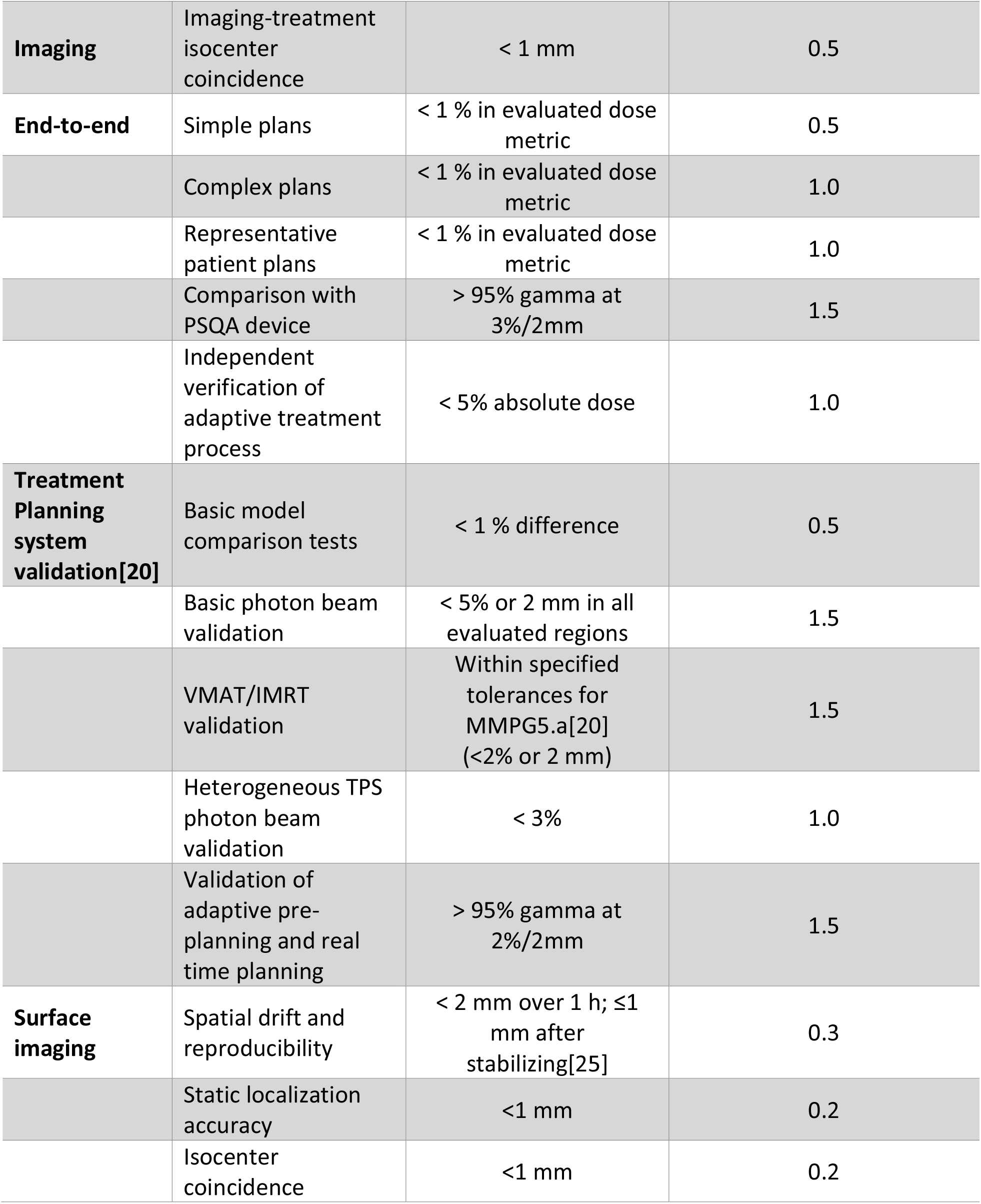
Summary of selected results from commissioning with an estimated time for completion

#### 3.1.4. Training timeline and resources needed

Clinical programs such as oART require input from all stakeholders within the radiation oncology team, and communication and concise decision making from within a defined group are crucial to successful implementation. To facilitate prompt and comprehensive overview of the oART program, a steering committee was established with a defined charge, temporal goals (both long and short term) and a governance structure to allow implementation and clinical integration. This committee consisted of members representing physicians, physicists, administration, therapy, dosimetry and nursing. This committee oversees the programmatic aspects of the oART program by providing guidance, training and constant evaluations of the direction and quality of the program.

User-specific training was carried out through a combination of vendor-led courses, clinical implementation/evaluation, and disease site-specific credentialing. Separate and defined training times and resources were used for each of the following groups: therapy, physics, dosimetry, physicians, and administrative/billing. Specific tasks and timelines for each of these groups are summarized in Table 3. User-specific training can be conducted in parallel to expedite clinical implementation. The Ethos treatment delivery application and planning system work independently from Aria and Eclipse, so appropriate time should be dedicated for all users to become comfortable with treatment planning, treatment delivery, and quality assurance workflows. The pre-acquisition evaluation time, prior to the purchase of the system, was not adequately reflected in Table 3 or Figure 1 but it is noteworthy and teams should prepare for accordingly. Furthermore, pre-implementation decision-making discussions (including workflow considerations, desired level of integration with existing systems, establishment of guidelines/procedures, and creation of staffing/coverage models) can be extensive and should include input from all members of the radiotherapy team. Therefore, adequate time should be set aside for these discussions.

**Table 3:** Breakdown of group specific training provided for the implementation of an oART program.

| Group | Training Objective | Estimated time (Days) |
| --- | --- | --- |
| **Therapy** | Basics of machine operation | 1-2 |
|  | Warmup and Daily QA | 0.5 |
|  | Primary patient set up and treatment of non-adaptive plans | 0.5 |
|  | Operational use of the surface imaging system | 1.5 |
|  | Setup and treatment of adaptive plans | 1.5 |
|  | Basic operation of Ethos treatment management software | 0.5 |
| **Physics** | Basics of machine operation | 1-2 |
|  | Advanced machine operation with service, QA and file modes | 1 |
|  | Patient specific quality assurance procedure | 0.5 |
|  | Ethos System Manager and shared framework portal | 1 |
|  | Plan creation, patient navigation, clinical goals, clinical priorities and contouring within ETM | 2 |
|  | Adaptive treatment operations and procedural training | 3-4 |
|  | Operational use of the Surface imaging system | 1.5 |
|  | Understanding Mobius3D Adapt | 0.5 |
| **Dosimetry** | Importing patients and setting up Intents in ETM | 0.5 |
|  | Plan creation, patient navigation, clinical goals, clinical priorities and contouring within ETM | 4-5 |
|  | Exporting/importing and file transfer from the TPS to ethos | 0.5 |
|  | Non-adaptive plan creation | 0.5 |
|  | Basics of adaptive sessions | 1 |
| **Physicians** | Basic operation of Ethos Treatment Management | 0.5 |
|  | Contouring, clinical goals, plan review, and dose monitoring | 1.5 |
|  | Adaptive treatment operations and procedural training | 1 |
| **Administrative/billing** | Changes to scheduling and departmental changes for adaption | 0.5 |
|  | Basic operation of Ethos Treatment Management | 0.5 |

### 3.2. Patient specific and ongoing quality assurance

Patient specific quality assurance (PSQA) was performed on the approved initial plans, also denoted as the scheduled plan, according to our department policy, with the ArcCheck (Sun Nuclear Corporation, Florida, USA) and Mobius3D-Adapt (Varian Medical Systems, Palo Alto, CA). Measurement passing criteria followed our department’s procedure for PSQA: 95% gamma[26] threshold using 3%/2mm criteria with exclusion of the 10% low dose region for ArcCheck, and 95% gamma threshold using 5%/3mm criteria for Mobius3DAdapt. Initially, we also performed a post-delivery PSQA verification on the first 20 generated adaptive plans and continued, for three months, with randomly selected adaptive plans for consistency verification. Detailed results from this post-delivery PSQA analysis can be found in the study by Zhao et al [27]. In summary, they analyzed randomly selected adaptive plans utilizing multiple PSQA methodologies and found that PSQA may not be necessary for verification of every adaptive treatment. Table 4 shows a summary of the ongoing quality assurance performed daily, monthly and annually.

**Table 4:** Summary of ongoing QA and required amount of time

| Category | Test | Equipment | Estimated time (Min) |
| --- | --- | --- | --- |
| **Daily** |  |  |  |
| **Safety** | Functional Interlocks and safety monitors | NA | 10 |
| **Dosimetric** | Output constancy | Daily QA Device | 10 |
| **Mechanical** | Virtual isocenter to machine isocenter verification | Hidden target phantom | 10 |
| **Mechanical/Imaging** | Phantom localization and repositioning | Hidden target phantom | 10 |
| **Monthly** |  |  |  |
| **Safety** | Functional Interlocks and safety monitors | NA | 10 |
| **Dosimetric** | Output constancy | Solid water with calibrated ion chamber and calibrated electrometer | 15 |
|  | Beam profile constancy | 2D detector array capable of measuring profiles | 15 |
| **Mechanical** | Virtual isocenter to machine isocenter verification | Hidden target phantom | 10 |
|  | Phantom localization and repositioning | Hidden target phantom | 10 |
|  | MLC positional and travel verification | NA | 10 |
|  | Winston Lutz test | Winston Lutz phantom | 10 |
| **Imaging** | Evaluation of Scale, distance, orientation accuracy, uniformity, noise, high contrast spatial resolution and low contrast spatial resolution | kV-CBCT specific Imaging Phantom | 10 |
| **Annual** |  |  |  |
| **Safety** | Functional Interlocks and safety monitors | NA | 10 |
| **Dosimetric** | Output constancy/ TG- 51 | 1D water tank with Calibrated Ion Chamber and Calibrated Electrometer | 30-60 |
|  | Profile constancy | 2D detector array capable of measuring profiles | 30-60 |
|  | "Flatness" and symmetry evaluation | 2D detector array capable of measuring profiles | 30-60 |
|  | Output factors | 1D water tank with Calibrated Ion Chamber and Calibrated Electrometer | 30-60 |
|  | MU linearity and PDD verification | 1D water tank with Calibrated Ion Chamber and Calibrated Electrometer | 30-60 |
|  | Independent verification of output | independent verification phantoms | 15 |
| **Mechanical** | Couch positional indicators | Ruler and graph paper | 10 |
|  | Virtual isocenter to machine isocenter verification | Hidden target phantom | 10 |
|  | Phantom localization and repositioning | Hidden target phantom | 10 |
|  | MLC positional and travel verification | NA | 10 |
|  | Winston Lutz test | Winston Lutz phantom | 10 |
|  | Collimator rotation isocenter | Film and Solid Water | 10 |
|  | Gantry rotation isocenter | Film and Solid Water | 10 |
|  | Table top sag |  | 10 |
| **Imaging** | Evaluation of scale, distance, orientation accuracy, uniformity, noise, high contrast spatial resolution and low contrast spatial resolution | kV-CBCT specific Imaging Phantom | 10 |
|  | Imaging dose evaluation | CTDI phantom | 20 |
| **End-to-End** | End-to-End verification of adaptive workflow | Adaptive verification phantoms | 60-90 |

### 3.3. Treatment

#### 3.3.1. Treatment Planning

Figure 2 and Figure 3 provide process maps for the adaptive and non-adaptive planning and treatment processes, respectively. For both adaptive and non-adaptive treatments physician target and normal tissue contouring was completed in Eclipse. For adaptive treatment planning, all planning was done within the Ethos treatment planning system. Although non-adaptive planning can be done entirely in the Ethos treatment planning system, a majority (>90%) of non-adapted plans were made in Eclipse and transferred into the Ethos system to utilize pre-existing physician and dosimetry workflows. This was an individual clinical preference, and both adaptive and non-adaptive planning can be done entirely in the Ethos treatment planning system. In our design of the non-adaptive planning workflow, we found that utilizing Eclipse as the planning system of record better integrated into our current clinical practice. Primarily using Eclipse also eliminated the need for Ethos-specific training for all of our dosimetry team. The non-adaptive treatment workflow follows the same standard operating procedures as conventional linear accelerator-based treatments in our clinic.

**Figure 2:**
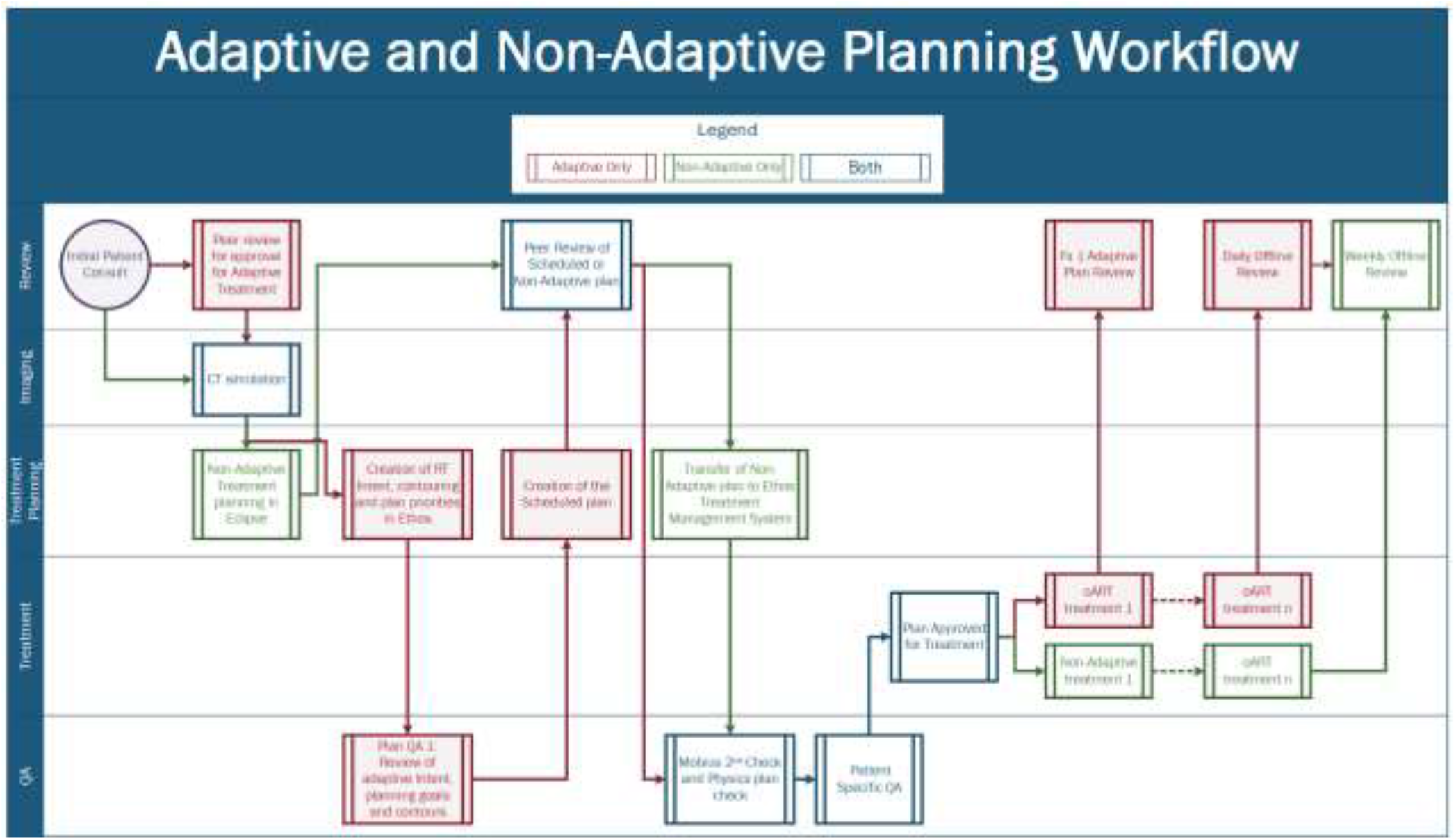
Adaptive and Non-Adaptive Planning Workflows separated by task type.

**Figure 3:**
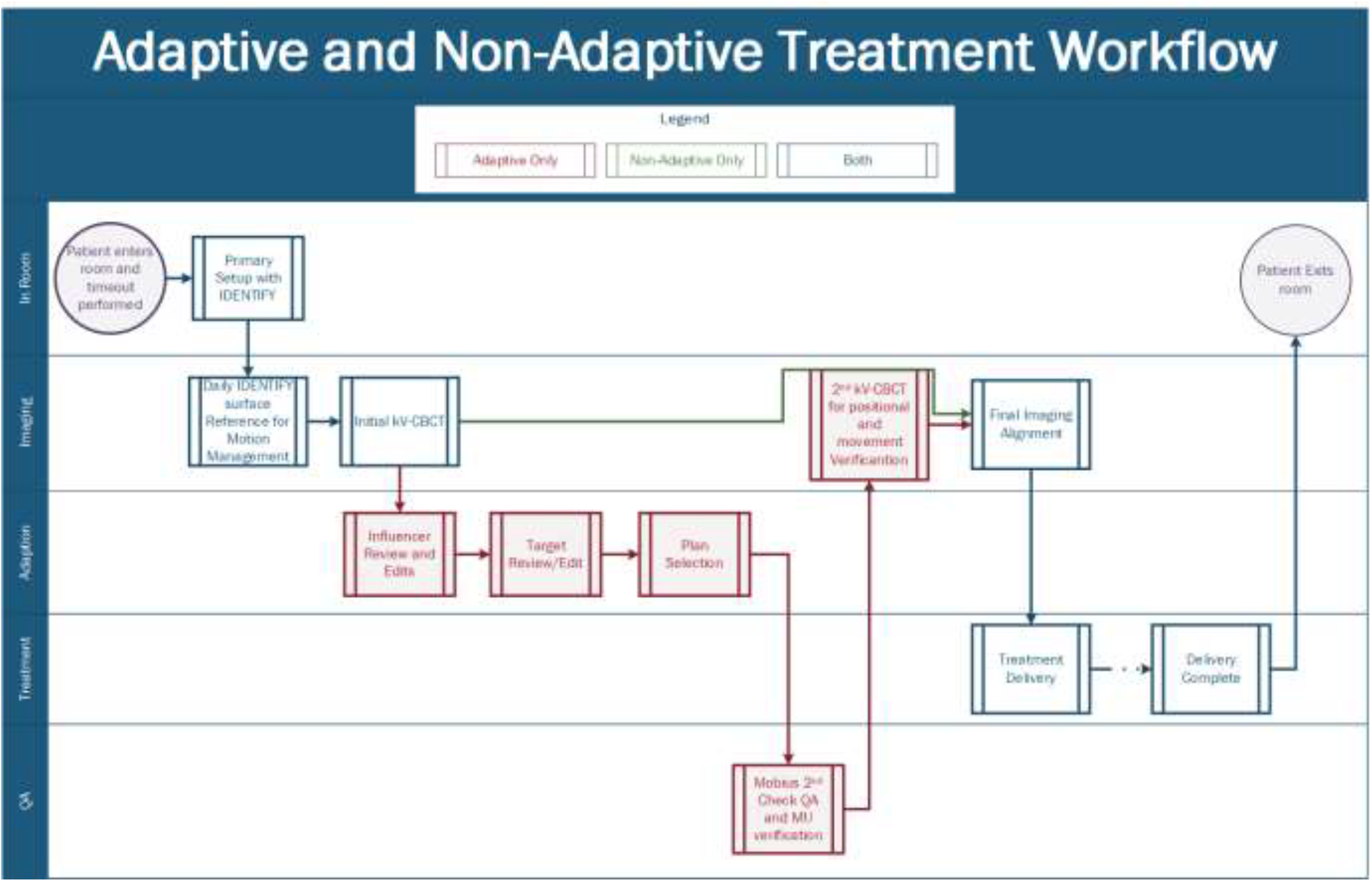
Adaptive and Non-Adaptive Treatment Workflows separated by task type.

For adaptive treatments, additional review and evaluation infrastructure had to be established to facilitate and ensure quality plan creation. Since the Ethos oART process starts with Influencer Structure review, these structures require special attention during the initial phase of planning. The review of contour derivation, influencers, margin generation, intent selection, priority goal ranking, planning image size and selected planning template became necessary steps to ensure high quality adaptive treatment sessions. Additionally, a significant amount of time was set aside to demonstrate the difference, between Eclipse TPS and Ethos TPS, for defined disease sites and specific plan types. Both systems rely on a grid-based Boltzmann solver for dose calculation [28, 29]. However, there are differences in the discretization of the CT image for dose calculation, leading to differences in reported DVH metrics. These differences between Eclipse-reported DVH and Ethos-reported DVH metrics should be understood. In our clinic, we chose to use Eclipse as the TPS of record. These differences, which are outside the scope of this report, were important to understand and highlight to facilitate comfort with the Ethos planning system.

#### 3.3.2. Treatment delivery

##### 3.3.2.1. Treatment workflows

The non-adaptive treatment delivery process, outlined in Figure 3, follows the traditional treatment process of primary patient setup, alignment with radiographic imaging, and delivery, consistent with conventional C-arm linear accelerators, with typically only the 2-3 treating therapists present. Also, kV-CBCT is the only imaging modality available on the Ethos system and is required for every patient. Additionally, surface imaging, with the Varian IDENTIFY, was utilized for primary positioning and intrafractional motion management. The IDENTIFY (Varian Medical Systems, Palo Alto, CA) is an optical surface imaging (SI) system used to monitor interfractional and intrafractional motion for all patients. Similar to other SI systems, IDENTIFY consists of three ceiling mounted camera pods, each containing two cameras and one projector. IDENTIFY uses a reference surface captured at the time of treatment for positional alignment and motion management throughout the course of treatment.[30] A region of interest (ROI) is selected for monitoring and its position is compared with the reference surface during treatment to monitor the patient’s overall movement. Differences in six degrees-of freedom, including longitudinal, lateral, vertical, pitch, roll, and yaw, are reported and monitored. During Delivery, if an offset larger than the defined thresholds is detected, treatment is stopped and another kV-CBCT is acquired.

The workflow for adaptive treatments is outlined in Figure 3. Clinical coverage consisted of a physicist, 2 therapists and the attending physician, when applicable. Per our institutional policy, physician presence was mandatory for the first adaptive fraction for conventional doses and once a week for subsequent fractions. For stereotactic treatments, physician presence is required for all fractions. A physicist was required to be present for the entirety of every adaptive treatment and performed all steps of the adaption process including, influencer edits and review, target edits and review, plan selection, and imaging verification. Following the adaptive workflow a 2^nd^ kV-CBCT verification image is acquired for every patient to verify final treatment position and evaluate temporal changes in internal anatomy.

The adaptive portion of the treatment delivery process is shown in Figure 4.This contains the steps outlined in the methods section above as well as specific processes implemented in our clinic, namely IDENTIFY for patient setup, Mobius for second check before treatment, and positional verification CBCT. During influencer review, the adaptor, a physicist in our case reviews the generated structures and edits contours as needed. To date, we have edited at least one contour for 100% of adaptive cases. Following influencer review, edits to the propagated target structures are commonly needed. These are typically minor, but it is not uncommon to need to entirely delete a target structure and re-contour from scratch, especially in the case of large anatomical changes such as tumor shrinkage. Furthermore, only influencers structures can be reviewed and edited prior to target generation, and since they guide sCT generation, special attention should be paid to these structures. Other structures (i.e. non-influencer structures) can be edited within the target review contouring workspace. Side by side comparison of the defined clinical objectives, shown in Figure 4C (orange is the scheduled plan, red is the adaptive plan), combined with the 3D dose distribution, allows for qualitative and quantitative plan evaluation. The reference plan, shown as the dashed line in the DVH of Figure 4C, represents the initial treatment plan that was approved at the time of planning on the original approved CT volumes, used for comparison at the time of plan selection. The adaptor then reviews the treatment plans, evaluating the scorecard on the left, dose distribution and DVH (seen in Figure 4). Generally, there is one plan which is superior for multiple DVH metrics and dose distribution. However, if the plans are quantitatively similar, treatment will be guided by patient-specific details, defined by the physician during planning and the first initial fraction. For more detailed information see references [31] and[32].

**Figure 4:**
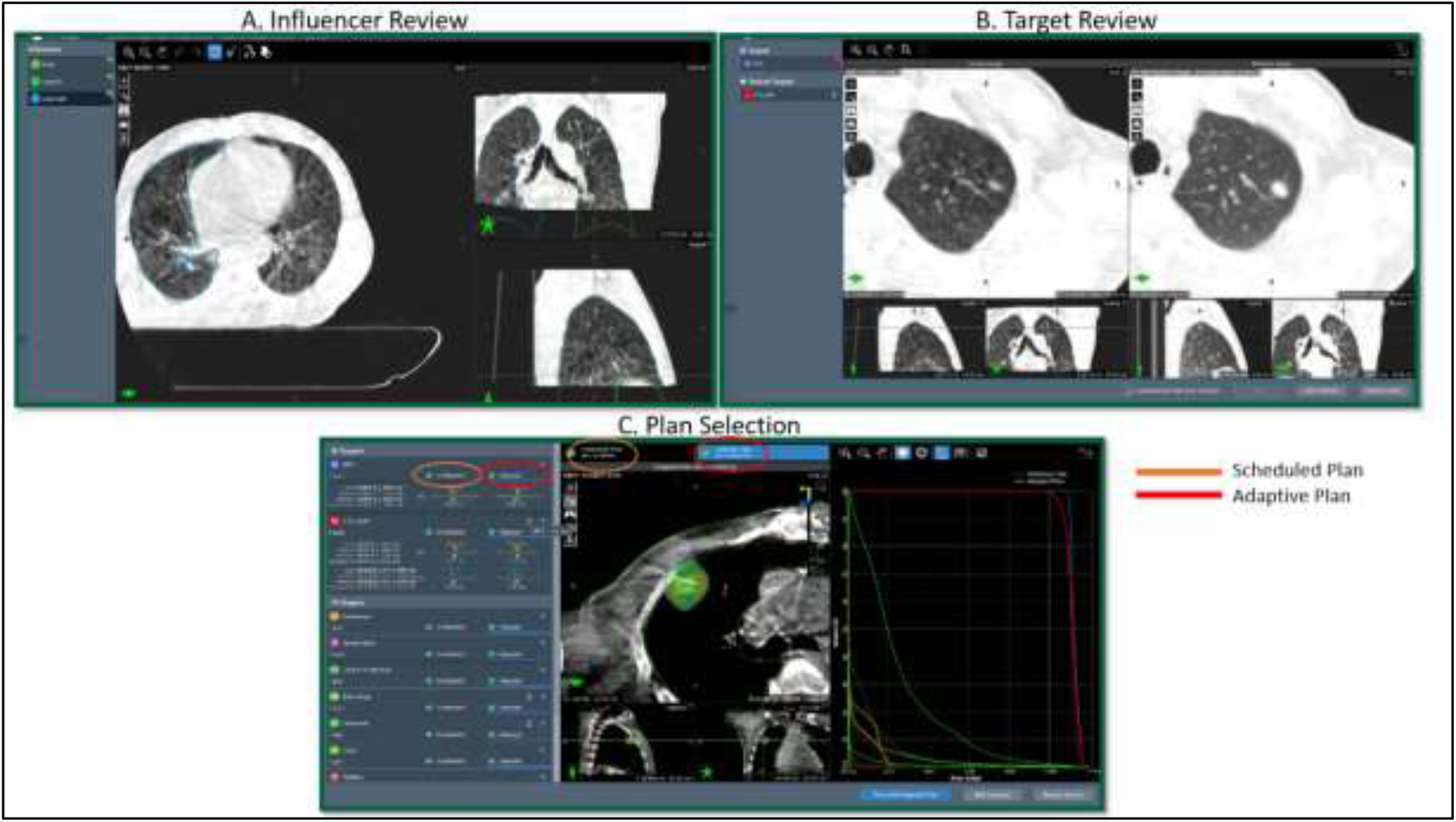
The adaptive portion of the treatment delivery workflow modules: A) Influencer Review, B) Target Review and C) Plan Selection with the adaptive plan visible.

As part of the adaptive workflow, all calculated plans’ DICOM files are automatically sent (after calculation is completed) to the Mobius3D Adapt verification for secondary dose verification, 3D gamma analysis, and DVH constraint assessment. MU verification is performed, between Mobius3D and the treatment machine, by the adaptive physicist to confirm that the correct MUs were transferred to the machine prior to treatment delivery. Following approval of the secondary calculation, a second CBCT, considered optional by the Ethos but required per institution policy, is acquired. This positional verification CBCT, used to correct for a residual patient movement and verify internal anatomy, is performed for every patient. It should be noted that this step is optional but highly recommended. Following delivery Mobius3D uses the machine trajectory log files to re-calculate the delivered dose as a post delivery assessment.

##### 3.3.2.2. Patient demographics and site breakdown

We retrospectively analyzed data from ninety-seven (N=97) patients treated from August 2021 through August 2022 for this UAB Institutional Review Board (IRB-1207033005) approved study. Table 5 shows a summary of the demographic information of the evaluated treated patients. Figure 5 shows the intent and fraction specific breakdowns of patients treated during this time. Patients were stratified into the following general categories based on general anatomical location of primary target and intent: Female Pelvis, Male Pelvis, Thorax, Breast, Abdomen, Head and Neck, and Other. For these 97 patients, a total of 1677 individual fractions were treated and analyzed. Figure 5 shows the site-specific breakdown of fractions treated during this time period separated by intent. 632 (38%) fractions were non-adaptive and 1045 (62%) were adaptive. 74 patients (76%) were treated with standard fractionation and 23 (24%) received stereotactic treatments. For the adaptive treatments, the generated adaptive plan was selected in 92% of treatments. The average age across all patients was 64 years (range 32-83); fifty-nine percent were male, forty-one percent female. Twenty-three percent of fractions were stereotactic doses. For the treatment delivery technique, 90% of the delivered fractions were intensity-modulated radiation therapy (IMRT) and 10% were volumetric modulated arc therapy (VMAT). Ninety-five percent of adaptive fractions were delivered using IMRT. This is primarily due to the increased optimization and calculation time needed for VMAT techniques with adaptive deliveries. Other groups have found enhanced plan quality with IMRT as compared to VMAT [33-35].

**Table 5:**
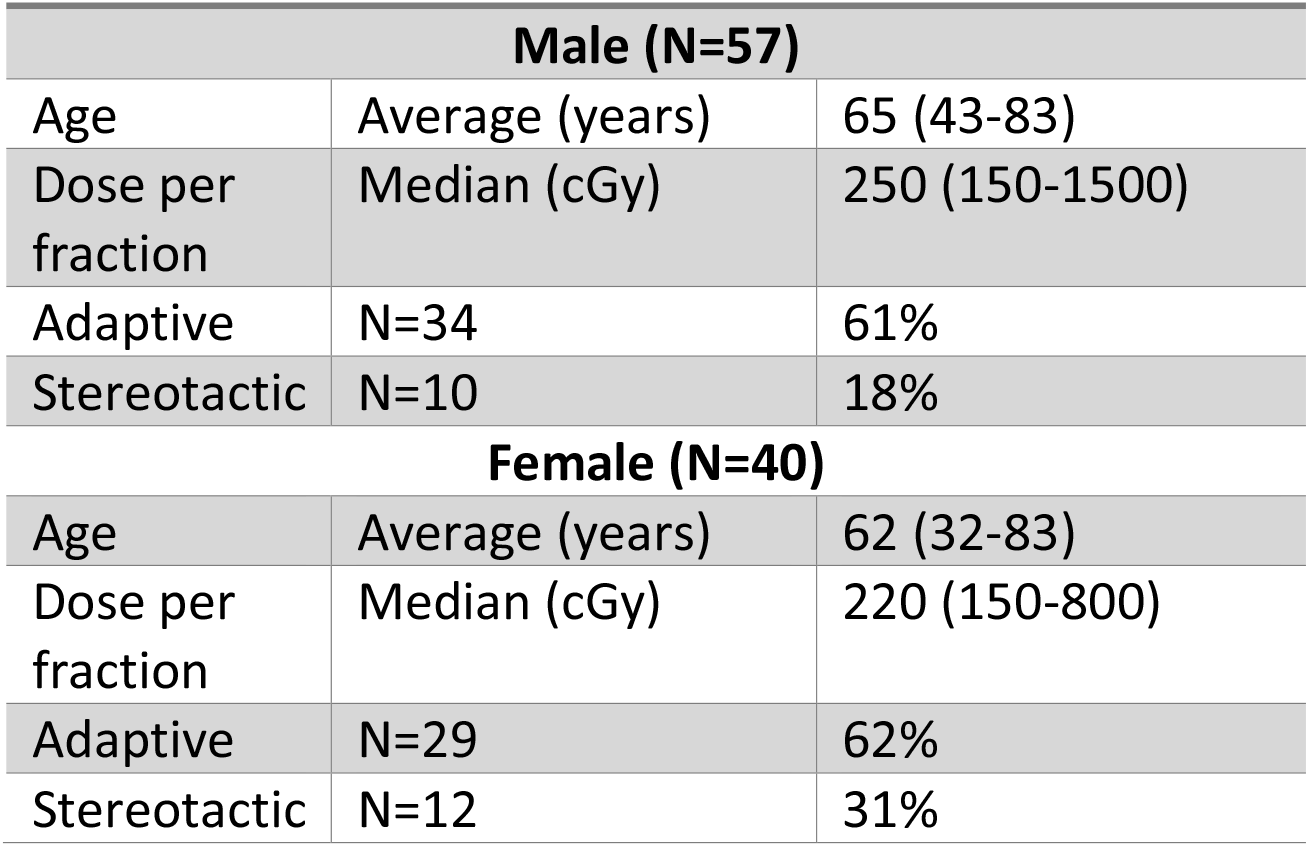
Demographic breakdown of patients treated

**Figure 5:**
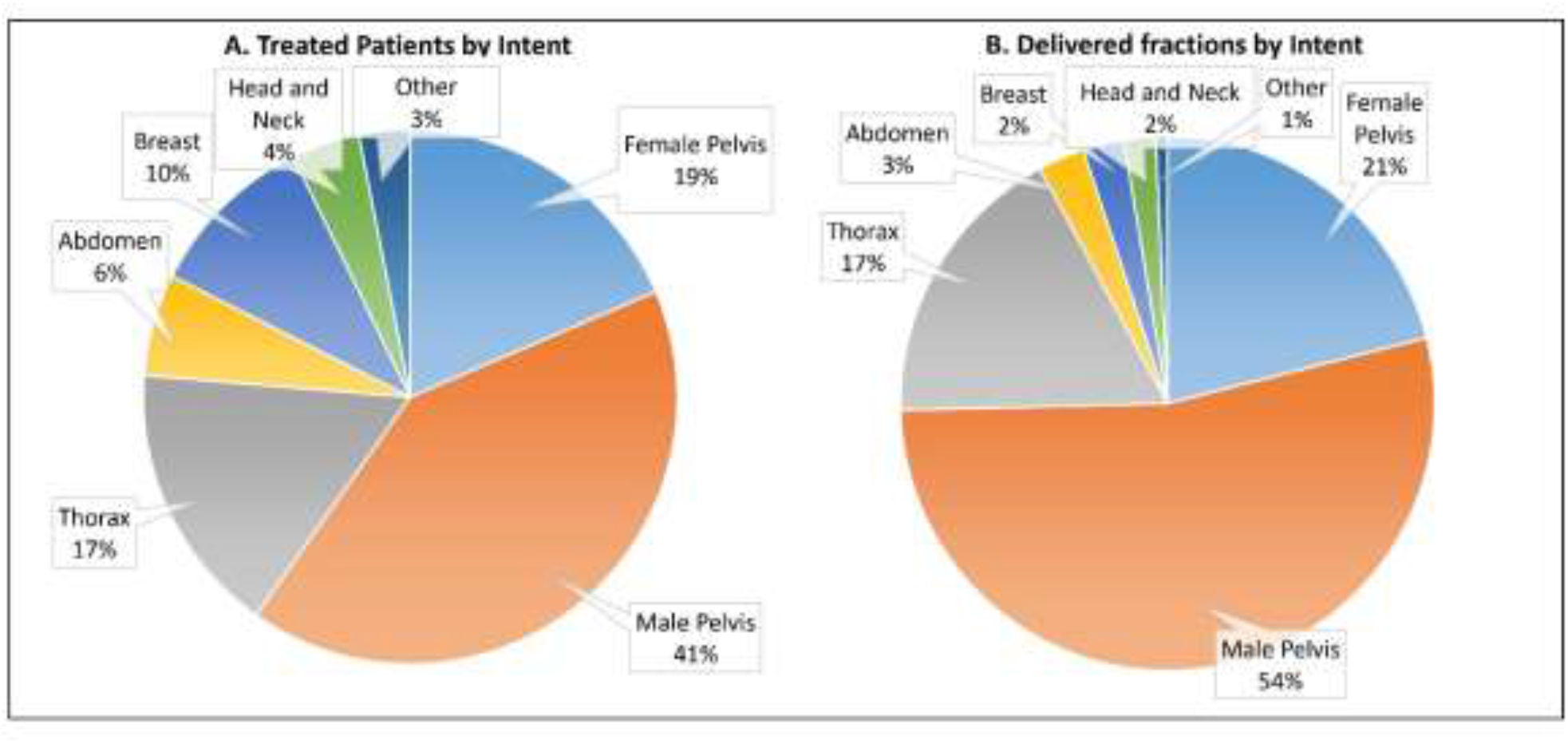
Representation of delivered fractions by A) patients per intent and B) delivered fractions per intent

##### 3.3.2.3. Analysis of treatment times

Table 6 and Figure 6 show the breakdown of treatment times by task for all of the analyzed fractions, separated into adaptive and non-adaptive treatments. On average (±std), adaptive treatments took 34.52 ± 11.42 minutes from start to finish. The average contour review and edit time for both the normal structures and targets was 9.50 ± 6.41 minutes. After contour approval, it took an average of 15.13± 6.15 minutes for plan generation, evaluation, and selection to start of treatment delivery. The entire adaptive process (from start of contour generation to the beginning of the verification CBCT), performed by the physicist (and physician on select days), was 19.84 ± 8.21 minutes. Mann-Whitney U unpaired, non-parametric test with significance level of 0.05 was used to determine that there was no difference in overall treatment time between standard-fractionated and SBRT adaptive treatments (p<0.05). Figure 7 shows the overall treatment time for all of the analyzed fractions separated into adaptive and non-adaptive treatments.

**Table 6:** The breakdown of treatment times by task for all of the analyzed fractions separated into adaptive and non-adaptive treatments

| **Adaptive** |  | Average<br>(min) | STD<br>(min) | Min<br>(min) | Max<br>(min) | Median<br>[Interquartile<br>range]<br>(min) |
| --- | --- | --- | --- | --- | --- | --- |
|  | Primary Setup | 9.36 | 5.85 | 1.47 | 44.27 | 7.81 [5.50] |
|  | Initial kV-CBCT review and<br>Influencer Contouring | 4.72 | 4.42 | 1.02 | 56.47 | 3.43 [2.25] |
|  | Target Review and editing | 4.78 | 4.02 | 1.01 | 29.47 | 3.68 [1.92] |
|  | Plan review and selection | 5.80 | 3.12 | 1.12 | 22.68 | 4.92 [3.82] |
|  | Mobius Second check Review and<br>MU verification | 4.55 | 2.10 | 1.33 | 17.53 | 4.07 [3.12] |
|  | Beam on time | 5.32 | 3.97 | 1.95 | 58.92 | 4.65 [3.93] |
|  | Overall treatment time | 34.52 | 11.42 | 15.62 | 90.67 | 31.93 [26.51] |
| **Non-**<br>**Adaptive** |  |  |  |  |  |  |
|  | Primary Setup | 9.18 | 4.88 | 1.42 | 35.82 | 7.89 [6.05] |
|  | Initial kV-CBCT review and<br>alignment | 3.80 | 2.07 | 1.07 | 17.13 | 3.26 [2.51] |
|  | Beam on time | 3.29 | 1.71 | 1.42 | 15.87 | 2.75 [2.33] |
|  | Overall treatment time | 16.82 | 8.76 | 3.68 | 52.57 | 14.18 [11.01] |

**Figure 6:**
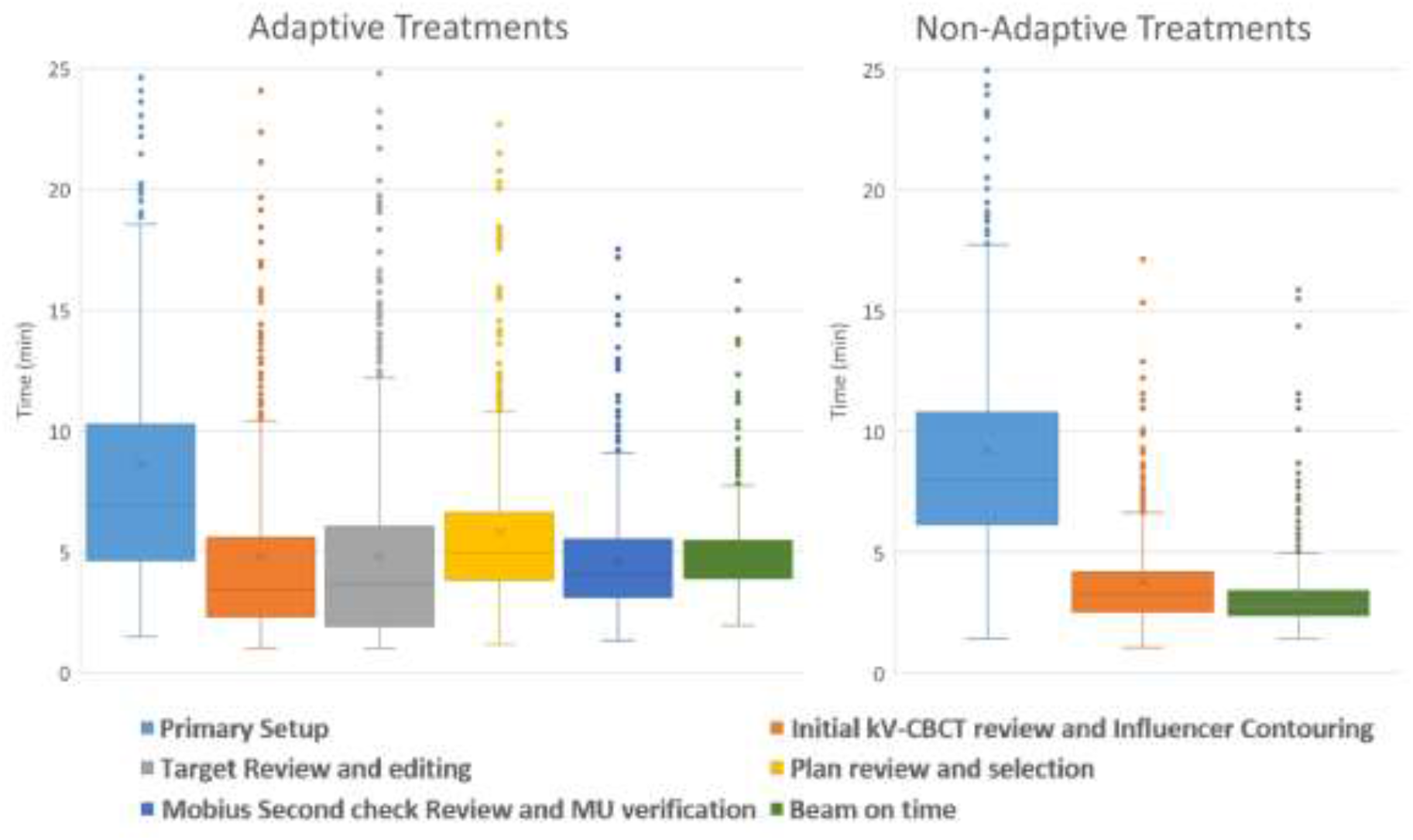
The breakdown of treatment times by task for all analyzed fractions, separated into adaptive and non-adaptive treatments. Outliers greater than 25 minutes are not show but are still included in the data analysis.

**Figure 7:**
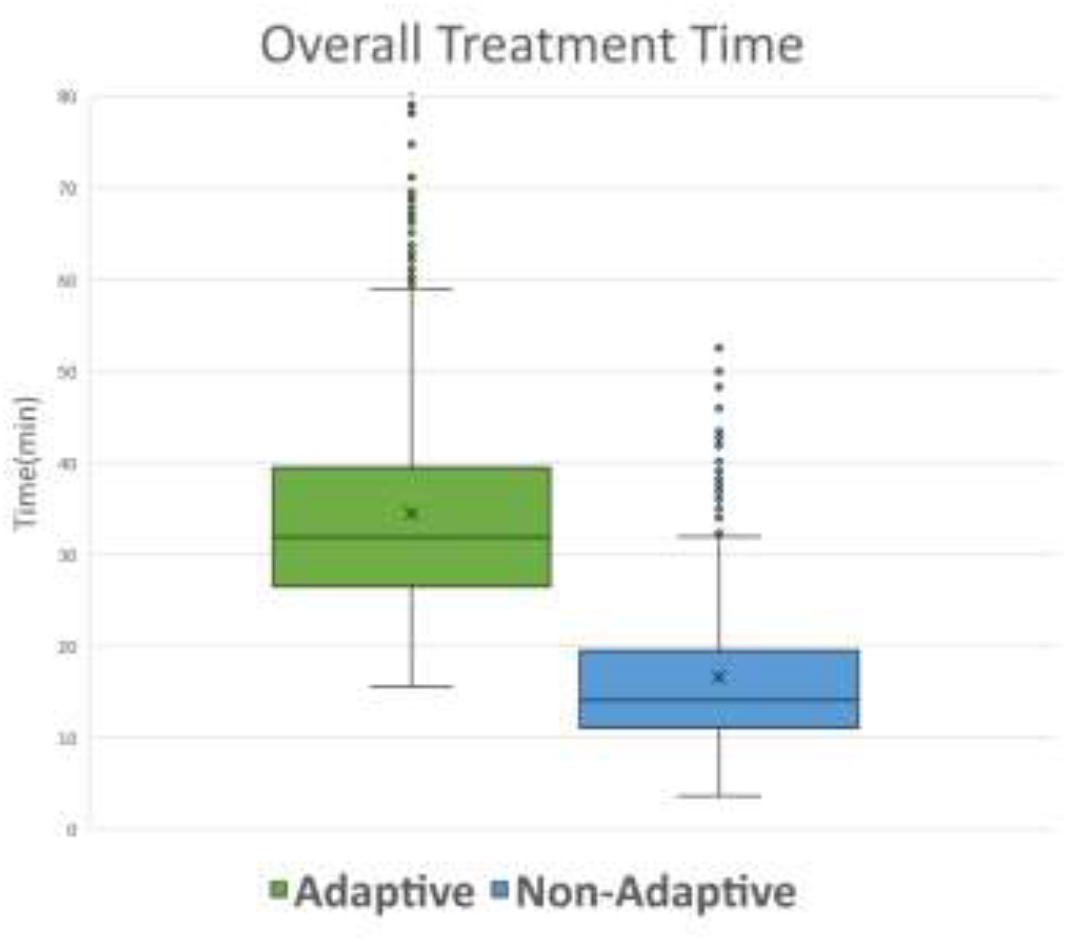
The breakdown of overall treatment time for all analyzed fractions, separated into adaptive and non-adaptive treatments. Outliers greater than 80 minutes are not shown but are still included in the data analysis.

## 4. Discussion

In this work, we present our institutions’ online adaptive radiation therapy program from pre-implementation stages to its current state. Furthermore, we provide, to our knowledge, the first retrospective time analysis describing our experience over the first year of our adaptive program using the Ethos treatment delivery system.

One of the major questions when starting an adaptive program is “how long does an adaptive treatment take?” As shown in Table 6, treatment times can vary significantly based on treatment site, complexity, patient anatomical specifics, number of structures and a multitude of other characteristics. The maximum treatment times for adaptive session, 89.87 and 90.67 mins, represent a prostate patient that urinated mid planning and a liver patient that moved significantly resulting is a session having to be restarted. In both instances, the entire adaptive process had to be restarted, contributing to the extensive treatment time. These outliers, while minimal in number, will contribute to a larger average treatment time but were included into the final analysis for the sake of transparency and clinical representativeness. Additional outliers can be seen for each individual step in the process and were also included. This highlights one of the most important factors of oART: selecting patients that will remain comfortable throughout the extended treatment times. Due to the lengthy adaptive process at the treatment console (not visible to the patient) it is imperative to select patients that would benefit from adaptive therapy for medical reasons and are also able to lie still for extended periods of time. Prolonged discomfort and inability to hold a position, lack of understanding, cooperation and claustrophobia have caused either significant treatment delays or cancellations of fractions entirely. Anecdotally, a thorough explanation of the treatment and expectations with the patients, prior to initial simulation and, again, prior to the first session have dramatically decreased the frequency of these cancellations. The fastest treatments, typically associated with the male pelvic patients, represented favorable anatomy that was clearly discernable and easily defined, with targets derived from the influencer structures and a small number of optimization objectives. Additionally, the majority of the longest treatment times represent VMAT plans. The cause for extended VMAT plan optimization time, an error in plan calculation and optimization that resulted in prolonged calculation times for “large” structures and OARs, was recently addressed in a software update. Even with this improvement, IMRT plans continue to optimize and calculate faster that VMAT plans and should be recognized and taken into account during treatment planning. Calculation times and time spent waiting are expected to be reduced by improvements in software upgrades, leading to a reduction of treatment time in the near future.

One of the most significant challenges that our group faced was the intensive amount of resources, both physical and operational, that an oART program requires. oART is a treatment technique that requires extensive understanding of internal anatomy, excellent communication skills, and is a skill that has to be practiced and maintained. Just as adaptive therapy is not suitable for all patients, it is not suitable for all physicists or physicians either. Special care should be paid when selecting members of an oART team to best position it for success. A considerable amount of time, prior to the installation of the machine, was spent on the training and evaluation of those involved with the oART program. Contouring evaluation, both with open source software[36] and one-on-one review with anatomical experts, was heavily utilized to not only demonstrate contouring efficiency but also instill a level of confidence with attending physicians regarding target evaluation and recognition. It is important to note that target delineation and modification were always verified by a qualified radiation oncologist prior to the next treatment. Additionally, oART treatments require a lot of hands-on time for the primary adaptors. While some people have adapted the model where the radiation therapist is the primary adaptor [37], we felt that this is outside the scope of a traditional therapist and should be done by a qualified medical physicist and/or medical doctor. This requires at least one qualified medical physicist, with dedicated and restricted clinical time, to be present at the machine for an average of 6 hours per day, and up to 8 hours, depending on the patient load. This should be taken into account when developing and evaluating staffing models for potential oART programs.

A significant limitation of the Ethos system was the lack of integration between the Ethos planning and delivery systems and Eclipse. Currently, Ethos only has the ability to pull basic demographic data, such as name, date of birth, and age, from eclipse but does not send any information back. This single sided information sharing has effectively turned ethos into an information “island” which resulted in altered clinical procedures and workflows, time-consuming workarounds and multiple duplications of effort to maintain our enterprise solutions. For example, after approving a plan in ethos, dosimetry will export the completed plan back into eclipse, where the prescription of record is located, and then manually import it and associate it to the patient. Then after each delivery, therapists complete a manual treatment in eclipse. This enables us to maintain dose tracking, weekly chart QA and plan summation capabilities; all of which are not present in the ethos environment. For large enterprise systems, this can serve as a significant barrier to implementation and its implications should be appropriately evaluated prior to implementation.

An additional technology, not directly evaluated in this study, that serves as an integral part of the oART process is the use of a surface imaging system, namely the Varian IDENTIFY. Surface imaging, while not required for oART, has become a crucial component of the workflow due to its ability to expedite patient position, monitor patient movement during the adaptive process and enable monitored breath-hold treatments. Throughout the lengthy treatments, it is not uncommon for patients to move. With the IDENTIFY, we have the ability to quantify the movement and use this information to provide intervention limits for repeat imaging and/or restarting the adaptive workflow. Research into the quantity and impact of these movements is underway. Utilization of automatic couch adjustments and gated adaptive deliveries are still under development by the vendor at the time of this manuscript, but will be quickly implemented into operational processes and workflows when available.

## 5. Conclusion

We present our institution’s experience establishing, organizing, and implementing an oART program using the Ethos therapy system. It took us 12 months from project inception to treatment of our first patient and 12 months to treat 1000 adaptive fractions. Retrospective analysis of delivered fractions showed that average overall treatment time was approximately 35 minutes and average time for the adaptive component of treatment was approximately 20 minutes. oART follows a different treatment paradigm than conventional RT and, because of this, the resources, implementation and workflows needed are more comprehensive and unique. Future studies will provide further guidance for radiation oncology teams to ensure the safe implementation of this new treatment modality.

## Data Availability

All data produced in the present study are available upon reasonable request to the authors

